# The Impact of Past Pandemics on Economic and Gender Inequalities

**DOI:** 10.1101/2021.04.28.21256239

**Authors:** Michal Brzezinski

## Abstract

This paper estimates how previous major pandemic events affected economic and gender inequalities in the short- to medium run. We consider the impact of six major pandemic episodes – H3N2 Flu (1968), SARS (2003), H1N1 Swine Flu (2009), MERS (2012), Ebola (2014), and Zika (2016) – on cross-country inequalities in a sample of up to 180 countries observed over 1950-2019. Results show that the past pandemics have moderately increased income inequality in the affected countries in the period of four to five years after the pandemic’s start. On the other hand, we do not find any robust negative impacts on wealth inequality. The results concerning gender inequality are less consistent, but we find some evidence of declining gender equality among the hardest hit countries, as well as of growing gender gaps in unemployment within the four years after the onset of the pandemic.

## 1. Introduction

The coronavirus SARS-CoV-2 pandemic has been a devastating health and economic shock. In 2020, there have been more than 80 million confirmed cases of COVID -19 including almost 1.8 million deaths. According to the IMF, global economic growth is projected to be −4.4% in 2020 (IMF, 2020). Although the growth is expected to rebound, the level of global GDP could be 3% lower by the end of 2021 than the level that would be obtained without the pandemic shock (Chudik et al., 2020). Moreover, health and economic disruptions have reduced household incomes and increased unemployment that could remain on an elevated level for an extended period. Several studies found that the pandemic hit particularly hard employment or working hours of young or precarious workers or those who work in alternative work arrangements and received relatively low wages already before the pandemic (Adams-Prassl et al., 2020; Crossley et al., 2021). Research established also that women were disproportionally affected by the crisis as it impacted more service sectors with high female employment shares and increased childcare needs during school closures (Alon et al., 2021; Sevilla and Smith, 2020). These developments could exacerbate existing economic and gender inequalities. Responding to this hypothesis, researchers started to study the distributional consequences of the past global health crises and the ongoing COVID -19 pandemic. With respect to the wage inequality effects of the present pandemic, Palomino et al. (2020) found that social distancing and lockdown measures increase the Gini coefficient in European countries in the range from 3.5% to 7.3%. Other studies also suggest that COVID-19 and short-term government policy responses could amplify existing wage and income inequalities (Almeida et al., 2020; Blundell et al., 2020). While the research on the distributional impact of COVID -19 is extremely important, it is also very difficult to estimate reliably the effects of the ongoing phenomenon. For this reason, a growing literature attempts to measure the impact of pandemics on inequalities by looking at past pandemics such as the 14th century Black Death, the 1918 Spanish Flu or the 1968 H3N2 Flu (Alfani, 2020; Furceri et al., 2020; Galletta and Giommoni, 2020; Saadi-Sedik and Xu, 2020).

The present paper contributes to the empirical literature on the effect of major past global health crises on inequalities by estimating the short- to medium-term impacts of recent pandemics on income, wealth and gender inequalities. ^1^ While some previous studies have already studied the impact of past pandemics on income inequality (Alfani, 2020; Furceri et al., 2020; Galletta and Giommoni, 2020; Saadi-Sedik and Xu, 2020), they have relied on income inequality measures computed from either household surveys or tax sources that either underrepresent top-income earners or do not cover the whole population (e.g. exclude non -tax-payers). On the other hand, the present paper uses newly-constructed, high-quality inequality data from the Distributional National Accounts (DINA) (Alvaredo et al., 2020b). The DINA provides cross-country comparable income inequality indices based on the combined data from household surveys, income tax sources and national accounts. By exploiting the DINA dataset covering 167 countries over 1950-2019, we aim at providing more reliable estimates of the impact of past pandemics on income inequality.

Additionally, to the best of our knowledge, this is the first paper to investigate econometrically how pandemics affected the distribution of wealth in the past.^2^ To this aim, we use wealth inequality measures for 46 countries observed over 2000 -2019 coming from Credit Suisse *Global wealth reports* (Davies et al., 2014). Finally, we study the empirical impact of past global health crises on gender inequality measured using composite and individual indices. The former indices include the UNDP’s Gender Inequality Index (GII) and the World Economic Forum’s Global Gender Gap Index (GGGI), while the latter use female unemployment rates and life expectancy measures. The data on composite gender inequality indices are available for more than 100 countries and cover the last three decades.

Section 2 of the reminder provides a short review of the literature measuring the impact of pandemics on various inequalities. In section 3, we present data and empirical methodology. Section 4 presents and discusses our results, while the last section concludes.

## 2. The impact of (past) pandemics on inequalities

### 2.1. Income and wealth inequalities

Alfani (2020) provides a detailed historical overview of the relationship between pandemics and income and wealth inequalities. He found that that the 14th century Black Death had an exceptional character in levelling wealth inequality. Later pandemic episodes until the cholera pandemics of the 19th century did not lead to sizable and lasting reductions in economic inequalities, while more recent global health crises like the 1918 Spanish flu seem to be rather inequality-increasing. This conclusion is confirmed by the study of Galletta and Giommoni (2020), who found that the 1918 pandemic permanently increased income inequality in Italy. De Santis (2020) has shown that the Spanish Flu increased income inequality across a group of countries.

Furceri et al. (2020) provide econometric evidence that five major recent pandemic episodes have increased income inequality (measured using survey -based data) in a panel of 175 countries over 1961-2017. They also found that pandemics significantly lowered the employment to population ratio for persons with basic education.

### 2.2. Gender inequality

There seems to be a lack of research on the impact of past pandemics on gender inequality. The existing studies focus on the case of COVID -19. For instance, Alon et al. (2020) argue that the pandemic is exposing mothers more than fathers to the risk of job loss as women work more frequently in occupations requiring face -to-face interactions or are unable to combine increased domestic responsibilities with demands of paid work. The least -educated mothers could be hit by the crisis particularly hard (Blundell et al., 2020). A related literature studies the impact of macroeconomic recessions on gender inequality. Doepke and Tertilt (2016) provide a summary of this research and show that women’s total labour supply is less volatile of the business cycle than men’s. Reasons for this fact include family insurance mechanism (with married women increasing their labour supply to compensate for higher unemployment risk of their husbands) and different sectoral composition of female versus male employment (with male -dominated sectors being usually much more significantly affected during the standard downturns). On the other hand, Seguino (2020) argues that prolonged recessions that lead to cuts in public sector budgets may have more negative impacts on women than men because women are over-represented in the public sector workforce.

## 3. Methods and data

### 3.1. Impulse response functions (IRFs) from local projections

Pandemic outbreaks are largely exogenous shocks to the economic systems. Their impact on inequalities and other outcomes can be estimated using impulse response functions (IRFs) – a standard macroeconomic tool capturing the dynamic response of a variable to the shock in another variable (see, e.g., Ramey, 2016). While there are several approaches to estimate the IRFs, the local projections (LP) m method of Jordà (2005) is simple, robust and provides straightforward inference tools (Olea and Plagborg-Møller, 2020).^3^ In essence, the LPs are linear regressions of the dependent variable shifted several periods ahead on the current and lagged values of the covariates. In our context, we estimate the following equations:

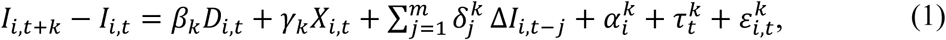

where *I*_*i,t*_ is the logarithm of an inequality index for country *i* in year *t, D*_*i,t*_ is a dummy variable indicating a pandemic shock occurring in the country *i* in year *t, X*_*i,t*_ is a vector of pre-determined control variables, Δ*I*_*i,t*−*j*_ are the lags of the change in the inequality index (with *m* set to 2).^4^ When we study the impact of pandemics on changes in income inequality, the control variables include the real GDP per capita and its square, population density, the urban population as per cent of the total population and the two KOF globalisation indices capturing globalization in trade and finance. The lags in the change of the (log of) inequality index are controlled for since future changes in inequality could depend on past changes. We also include country fixed effects, 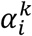, and time fixed effects, 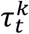, to control for unobserved, respectively, time –invariant and country-invariant heterogeneity (i.e. global economic shocks).

In our empirical analysis, we conduct separate estimations of IRFs for several indicators of income, wealth and gender inequalities presented in detail in section 3.2. The estimated *β*_*k*_ regression coefficients in (1) represent LP estimates of IRFs to the pandemic shocks happening at time *t*. They measure the percentage change in inequality in countries affected by the pandemics relative to the unaffected countries. The responses of inequality measures are displayed on figures as the dynamics of 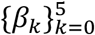 for the time horizons of up to five years after the shock. We also present 90% confidence intervals for our estimates calculated with standard errors clustered at the country level. Full regression results are presented in Tables A3-A12 in the Appendix.

### 3.2. Data sources

#### 3.2.1. Pandemic events

We follow Ma et al. (2021) in focusing on the six postwar pandemics: H3N2 Flu (1968), SARS (2003), H1N1 Swine Flu (2009), MERS (20212), Ebola (2014), and Zika (2016).^5^ Table A1 in the Appendix presents the list of all countries affected during a given pandemic. Our main pandemic-related variable is a dummy marking the timing of the pandemic’s outbreak according to the official WHO declarations. However, since the impact of different pandemics varied significantly by country in terms of mortality due to the pandemic or other health outcomes we exploit also additional measures of crisis severity. We follow Ma et al. (2021) in using three dummy variables capturing the relative health severity of the pandemics. High -severity countries are defined as those that experienced pandemic mortality higher than 70th percentile of the distribution of cross-country mortality due to the pandemic. Medium-severity countries are those in which pandemic mortality was within the range between the 30th and 70th percentile of mortality distribution, while for low -severity countries the mortality was below the 30th percentile.

In the baseline sample covering the 1950 -2019 period, we have 294 country-year observations indicating the start of the pandemic shock.

#### 3.2.2. Income inequality data

We use inequality and redistribution data from the recently compiled novel Distributional National Accounts (DINA) that provide harmonized, cross -country comparable information on the distribution of country’s entire national income consistent with macroeconomic aggregates from the System of National Accounts (Alvaredo et al., 2020b, 2020a). Previous research on the impact of pandemics on income inequality relied on data from household surveys (Furceri et al., 2020). However, income data from household surveys significantly underrepresent top incomes and usually do not cover all sources of income. In consequence, estimates of levels and trends in survey-based inequality measures are sizeably downward biased. On the other hand, the DINA database offers inequality indices computed using the combined data from income tax sources, household surveys and national accounts (including tax -exempt capital income and undistributed profits) in a way that provides much more complete estimates of the income distribution in the whole population. Therefore, we expect that our estimates of the impact of pandemics on income inequality can be more reliable than those obtained in previous studies. Our main income inequality indices from DINA are top income shares and the Gini coefficient in for the pre -tax pre-transfer national income. ^6^ The sample covers 167 countries over the 1950-2019 period.

#### 3.2.2. Wealth inequality data

The wealth inequality data come from Credit Suisse *Global wealth reports*.^7^ The reports provide data on inequality in the distribution of net worth defined as the sum of households’ financial and non-financial assets minus debts. The primary wealth inequality indicators available are wealth shares for the top decile and the top 1% of wealth distribution. The 2014 Global wealth report provided top wealth shares for 46 countries over the period 2000 -2014 (Davies et al., 2014). We updated this information with annual top wealth shares available in 2015 -2019 editions of the *Global wealth reports*. The final sample covers 46 countries observed over 2000 - 2019.

#### 3.2.3. Gender inequality indicators

Regarding gender inequality indices we consider both composite measures and individual indices. The former group includes the UNDP’s Gender Inequality Index (GII) and the World Economic Forum’s Global Gender Gap Index (GGGI) (Stotsky et al., 2016). The former measure is based on three components including female reproductive health, women’s empowerment and their labour market status. Higher values of the GII indicate greater gender inequality. The GGGI is constructed using 14 separate indicators of gender gaps in four areas: educational attainment, health and well -being, economic participation and opportunity and political empowerment. The GGGI is increasing in gender equality. The data on GII cover 155 countries over 1995-2019 period, while the GGGI index is available for 133 countries and the 2006 -2020 period.

Beside composite indices, we rely also on some individual health and economic indicators that are more transparent and easier to interpret. In particular, we study changes in the female unemployment rate and female life expectancy, as well as their relative versions (ratio of female to male unemployment rate and ratio of female to male life expectancy). The data are taken from the World Bank’s World Development Indicator (178 countries over 1990-2020).

## 4. Results

### 4.1. The impact of recent pandemics on income and wealth inequalities

Figure 1 shows estimated IRFs based on LP, estimated using equation 1, for the effect of pandemic shocks on pre-tax pre-transfer income inequality measured using the top 1% income share and the Gini coefficient. ^8^ The black lines show the evolution of income inequality in countries affected by the six analysed pandemics relative to unaffected countries over the period of five years since the onset of the shock. Our estimates show that the inequality-increasing effects of health shocks start to be statistically significant after two years since the onset of the event and remain significant within the five-year (top 1% income share) or four-year time horizon. After four years, the top 1% income share are on average higher in the affected countries by 3.2% compared to the unaffected countries. This amounts to about 20% of the standard deviation of 4-year changes in the top 1% shares in the estimated sample. Therefore, the effect is rather moderate in size. The absolute effect in case of the Gini index is smaller (1.2% after four years) and becomes insignificant after five years. These results suggests that past pandemics had a larger impact on inequality at the top of income distribution than in the middle or at the bottom of it. It might be that incomes of the poor and middle-class households have been recovering from pandemic shocks in the medium term (five years) counteracting the earlier rise in a comprehensive inequality measure such as the Gini index. On the other hand, the income gains to the top-income earners have been more persistent leading to a more sustained higher level of top-income inequality.

**Figure 1.**
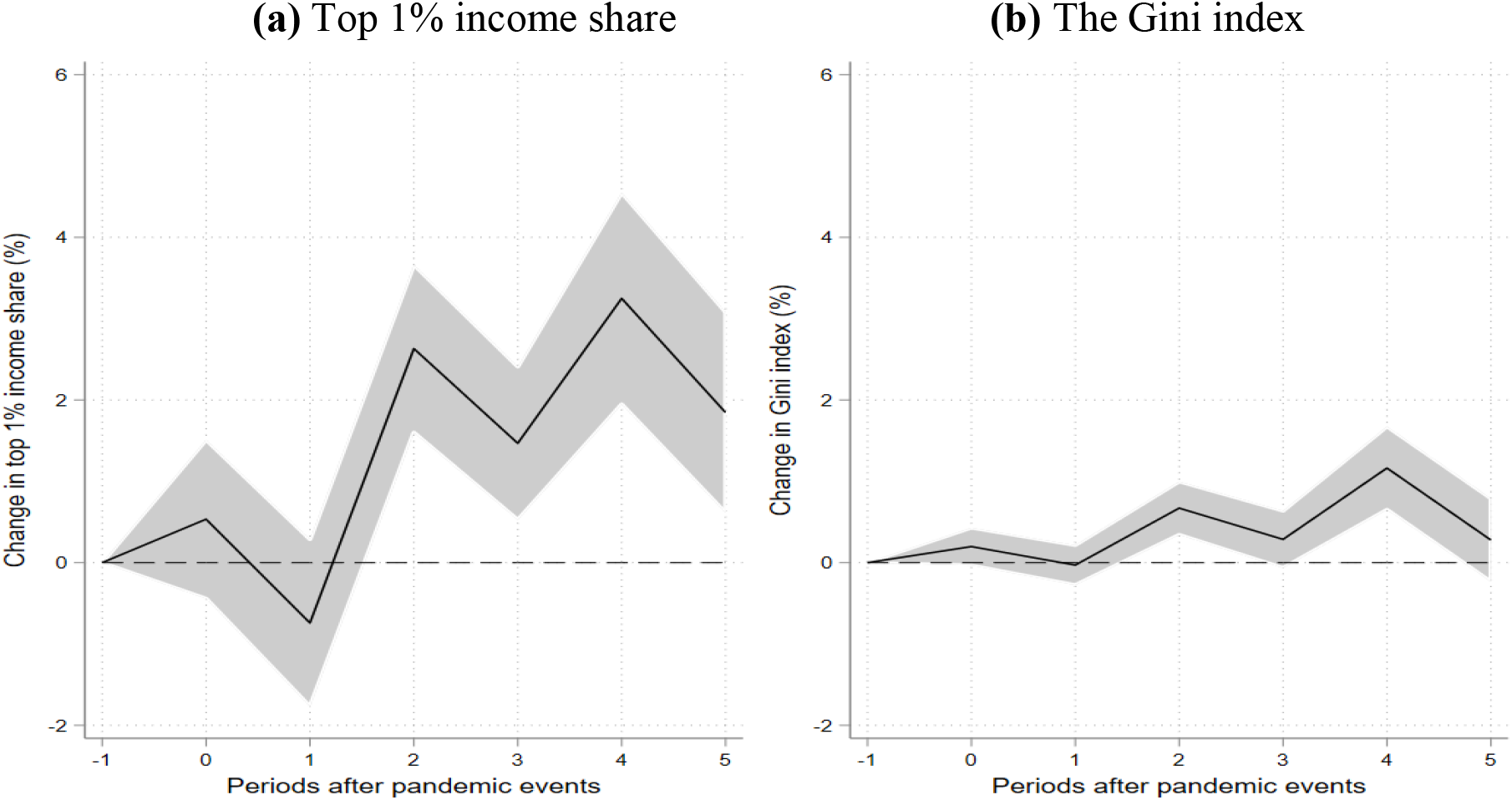
The response of income inequality to the shock from pandemic episodes. *Note*: Estimates of IFRs based on LPs (see equation 1). The dependent variable is the change in the logarithm of the top 1% income share (left panel) or change in the logarithm of the Gini index (right panel). See section 3.1 for a description of the model specification. Shaded areas are 90% confidence intervals. The horizontal axis shows years (*k*) after the start of the pandemic event with *k* = 0 denoting the year when the pandemic started. The sample covers 167 countries over the 1950-2019 period.

Our results for pre-tax pre-transfer inequality measures are broadly consistent with those of Furceri et al. (2020) obtained with survey-based inequality data. However, they found that the effect on the Gini for market incomes is marginally significant even in the five -year horizon. Another difference is that Furceri et al. (2020) found that the previous pandemic shocks had a greater impact on post-tax post-transfer inequality measures than on the pre-tax pre-transfer indices. Unfortunately, as for now the DINA database offers estimates of inequality measures for post-tax post-transfer incomes only for about 40 advanced countries for the period since 1980. Therefore, these data do not capture most of our pandemic episodes. However, even in this small sample the pandemics led to an almost 7% increase in the top 1% post-tax posttransfer income share within two years after the onset of the pandemic. The estimated effects are smaller for the top 10% income shares and insignificant for the Gini index.^9^

We did not find a statistically significant impact of past pandemics on average extreme income poverty rates measured using a variety of poverty lines. ^10^ This may indicate that in case of previous pandemics social safety nets (unemployment benefits, welfare payments and other cash or in-kind transfers) were in general strong enough to compensate income losses of the households at the bottom of the income distribution. On the other hand, the effect of pandemics on absolute poverty seems to be highly sensitive to the severity of the crisis. In particular, we found that in the high-severity countries absolute poverty increased by almost 20% four years after the onset of the pandemic. However, the effect becomes insignificant in the fifth year. This results suggests that the impact of the COVID-19 pandemic on poverty could be devastating, especially in the hardest hit countries. Simulations by the World Bank suggest that due to the pandemic the number of the world poor could increase by at least 131 million in 2020 (Yonzan, N., 2020).

Our results for the pre-tax pre-transfer top 1% income share that take into account differences in crises severity by country are reported in Figure 2. The effects are large and significant mainly for the high -severity countries. They are substantially smaller and rarely significant for the medium and low-severity countries. Overall, these results suggest that inequality-increasing effects of past pandemics occurred mainly in the countries that suffered the largest health cost in terms of pandemic-related mortality.

**Figure 2.**
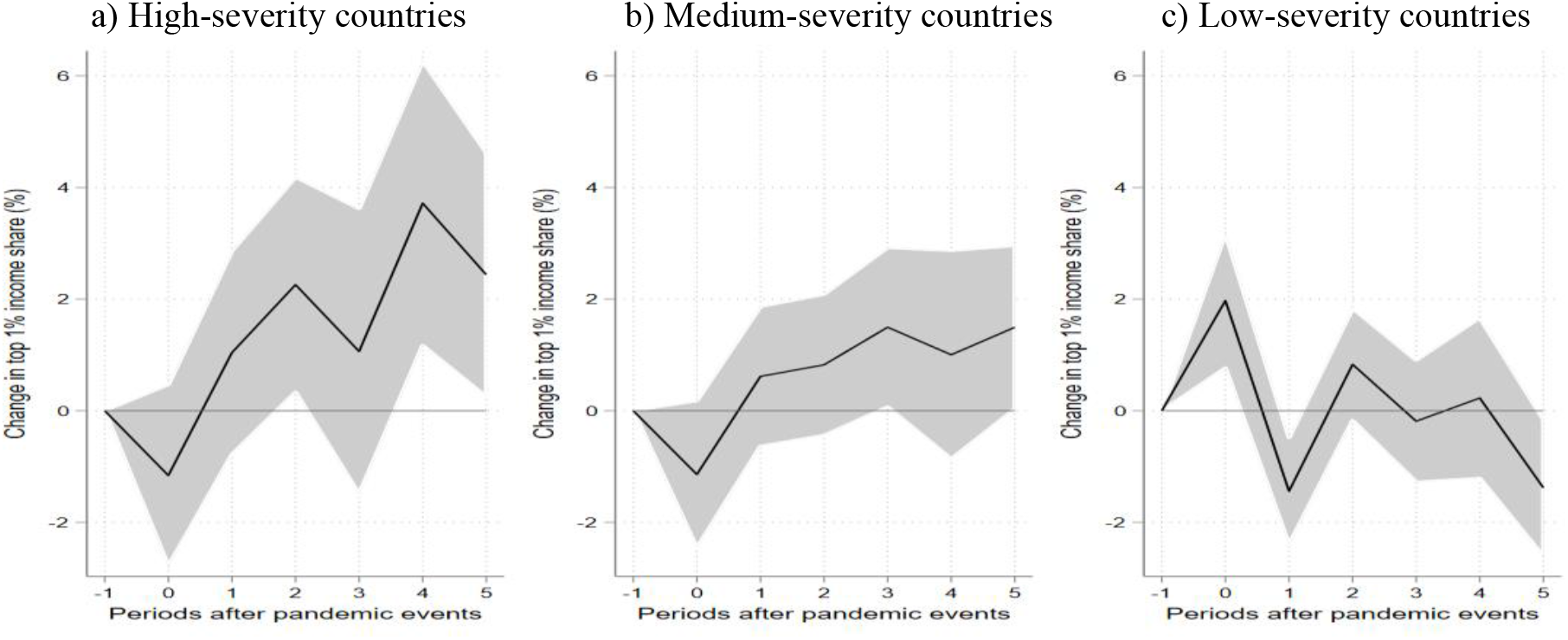
The response of income inequality to the shock from pandemic episodes by pandemic severity. *Note*: Estimates of IFRs based on LPs (see equation 1). The dependent variable is the change in the logarithm of the top 1% income share. See section 3.1 for a description of the model specification and section 3.2.1 for the definition of country groups by pandemic severity. Shaded areas are 90% confidence intervals. The horizontal axis shows years (*k*) after the start of the pandemic event with *k* = 0 denoting the year when the pandemic started.

Figure 3 presents results on the short- to medium-term impact of previous pandemics on wealth inequality measured using the top wealth shares. We do not find any statistically significant effects.^11^ We observe a 1% drop in the top 1% wealth share one year after the onset of pandemics, but this effect is not significantly different from zero. The impact of pandemics on wealth inequality is theoretically ambiguous. On the one hand, health and economic crises brought about by the pandemics could hurt employment and education opportunities for the poor and middle-class persons contributing to the rising wealth inequality in the longer run. On the other hand, the pandemic shocks could have a negative impact on stock markets and rates on return on assets. Such mechanisms should compress wealth distribution at the top and lower the overall wealth inequality. Indeed, Jordà et al. (2020) show that major past pandemics (ranging from the 14 century Black Death to the 2009 H1N1 pandemic) have depressed real rates of return on assets in the 40-years horizon after the start of a pandemic. Regarding the stock market channel, Baker et al. (2020) show that past pandemics did not have a substantial effect on the US equity markets. Overall, it might be that different pandemic -induced effects on wealth inequality cancel each other out or that the joint effect manifests itself over a longer time horizon than admissible in the present paper. Our results may be also explained by the fact that our sample is limited to only 46, mostly advanced, countries observed over a relatively short period (2000-2019).

**Figure 3.**
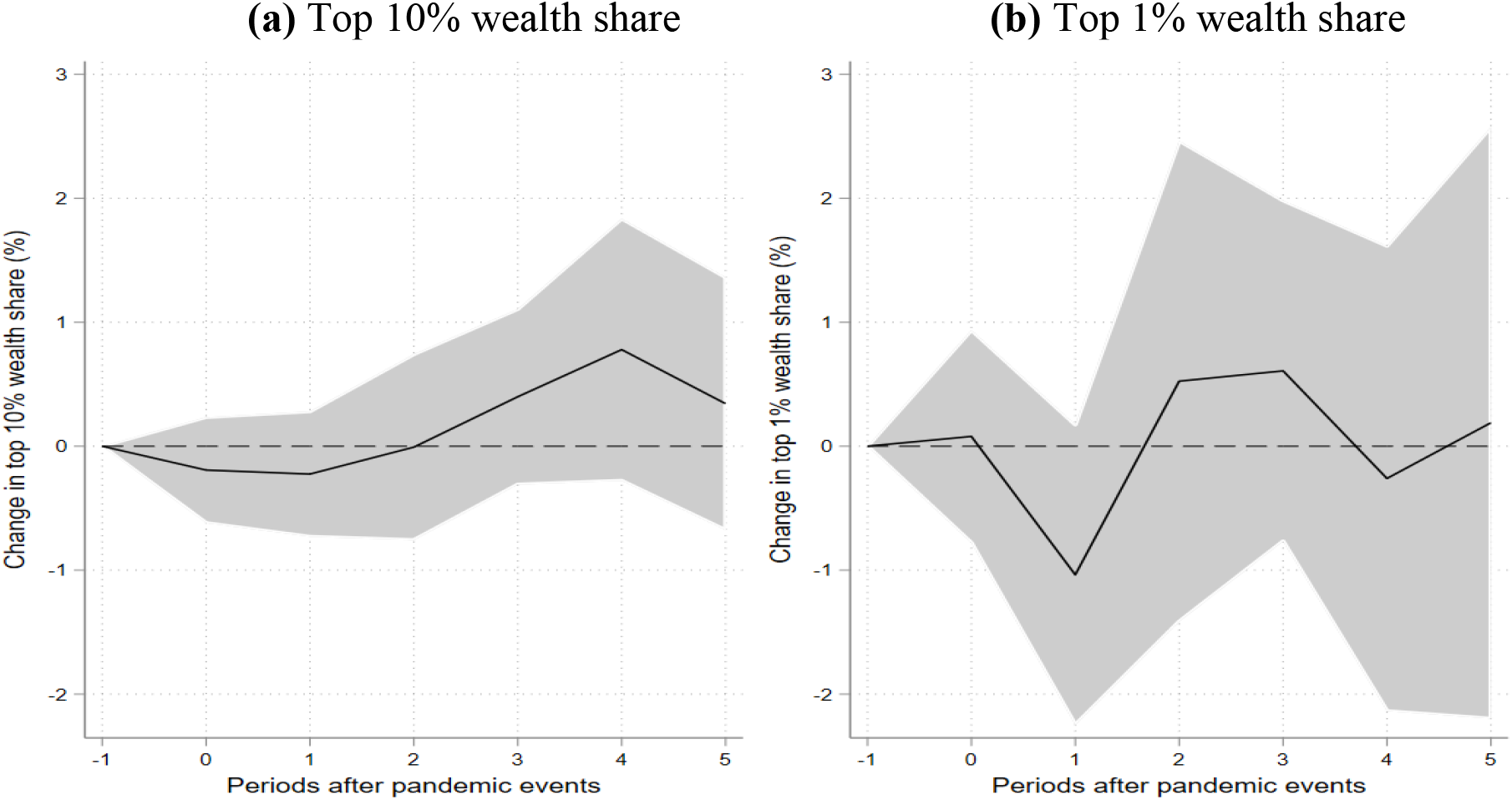
The response of wealth inequality to the shock from pandemic episodes. *Note*: Estimates of IFRs based on LPs (see equation 1). The dependent variable is the change in the logarithm of the wealth share held by the top 10% (left panel) or top 1% (right panel) of the population. See section 3.1 for a description of the model specification. Shaded areas are 90% confidence intervals. The horizontal axis shows years (*k*) after the start of the pandemic event with *k* = 0 denoting the year when the pandemic started. The sample covers 46 countries observed over 2000-2019.

### 4.2. Effects on gender inequality

Results for the impact of past pandemics on composite indices of gender inequality are presented in Figure 4. We observe a small (2.2%) increase in the GII three years after the pandemic’s onset. However, this positive effect quickly disappears in the fourth year after the health shock. Therefore, we do not find a robust impact of past pandemics on gender inequality measured using the GII. The effects for the GGGI do not show a clear pattern. A year after the pandemic onset there seems to be a small (0.8%) increase in gender equality, which, however, dissipates over the following three years and even turns negative (indicating decreasing gender equality) in the fifth year after the pandemic’s start. Further insights regarding the GGGI can be gained by looking at the estimates by pandemic severity displayed in Figure 5. ^12^ Interestingly, we observe a sizeable and significant reduction in gender equality in countries heavily affected by the pandemics that is sustained over five years. On the other hand, low -severity countries experience an increase in gender equality as measured by the GGGI which, however, disappears after four years. Overall, these results do not show a clear pattern of increasing gender inequalities due to the past pandemics over the five -year horizon. However, there is a significant negative impact of the pandemics on gender inequality when measured using the GGGI in the case of the hardest hit countries.

**Figure 4.**
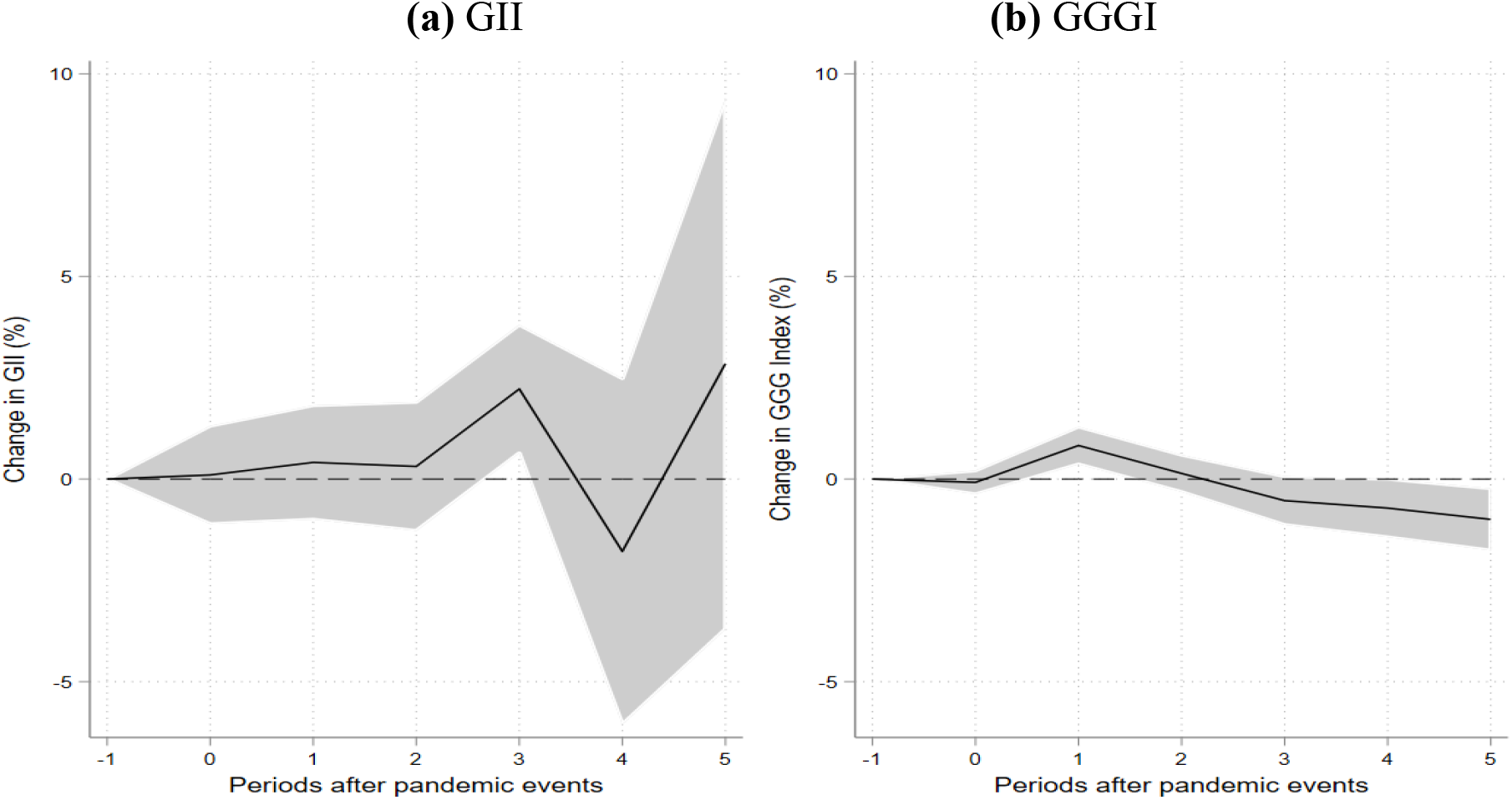
The response of gender inequality indices to the shock from pandemic episodes. *Note*: Estimates of IFRs based on LPs (see equation 1). The dependent variable is the change in the logarithm of the Gender Inequality Index (left panel) or change in the logarithm of the Global Gender Gap Index (right panel). See section 3.1 for a description of the model specification. Shaded areas are 90% confidence intervals. The horizontal axis shows years (*k*) after the start of the pandemic event with *k* = 0 denoting the year when the pandemic started. The sample for GII covers 155 countries over 1995 -2019 period, while that for the GGII – 133 countries over 2006-2020.

**Figure 5.**
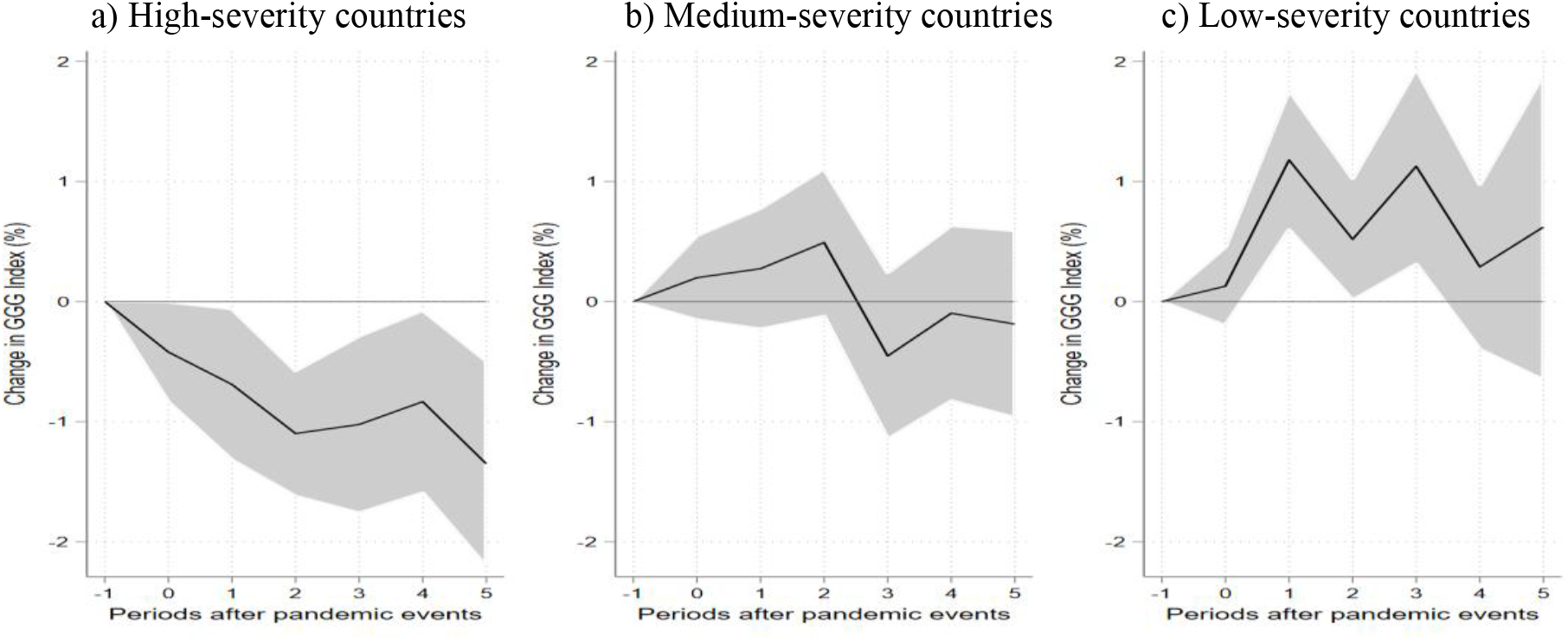
The response of the Global Gender Gap Index to the shock from pandemic episodes by pandemic severity. *Note*: Estimates of IFRs based on LPs (see equation 1). The dependent variable is the change in of the Global Gender Gap Index. See section 3.1 for a description of the model specification and section 3.2.1 for the definition of country groups by pandemic severity. Shaded areas are 90% confidence intervals. The horizontal axis shows years (*k*) after the start of the pandemic event with *k* = 0 denoting the year when the pandemic started.

Our estimates of the response of individual gender inequality indicators to the pandemic shocks are presented in Figures 6-7. Figure 6a shows that the contemporaneous effect of the health shock to female unemployment is relatively large (3.7%) and statistically significant. It is gradually reduced over time and falls to zero at the end of the five-year horizon. The relative measure depicted in Figure 6b shows that the change in female unemployment relative to male unemployment grows up to the fourth year after the pandemic’s start but then it falls to zero in the fifth year. This measure provides some support for growing gender gaps in unemployment, but the effect is small (3.7%) and not persistent.

**Figure 6.**
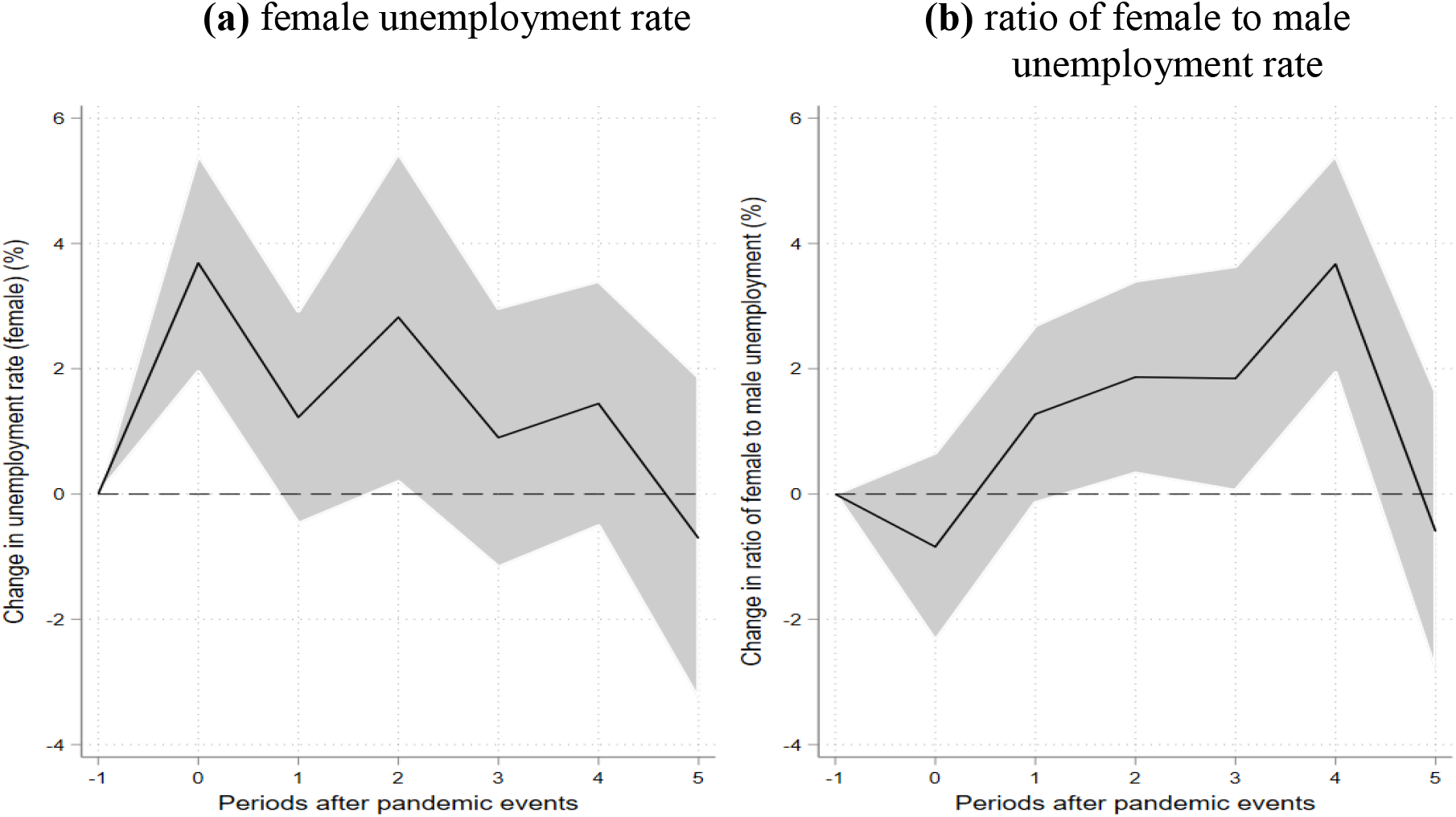
The response of unemployment indices to the shock from pandemic episodes. *Note*: Estimates of IFRs based on LPs (see equation 1). The dependent variable is the change in the logarithm of the female unemployment rate (left panel) or change in the ratio of female to the male unemployment rate (right panel). See section 3.1 for a description of the model specification. Shaded areas are 90% confidence intervals. The horizontal axis shows years (*k*) after the start of the pandemic event with *k* = 0 denoting the year when the pandemic started. The sample covers 178 countries over 1990-2020.

Figure 7 presents the results in terms of female life expectancy and ratio of female to male life expectancy. Female life expectancy drops persistently by about 0.1% over the five - year horizon (Figure 7a). However, gender inequality in terms of gendered life expectancy does not show significant changes (Figure 7b). Overall, there seems to be no evidence that the past pandemics have increased gender inequality in health measured by life expectancy in the period of five years after the pandemic’s start.

**Figure 7.**
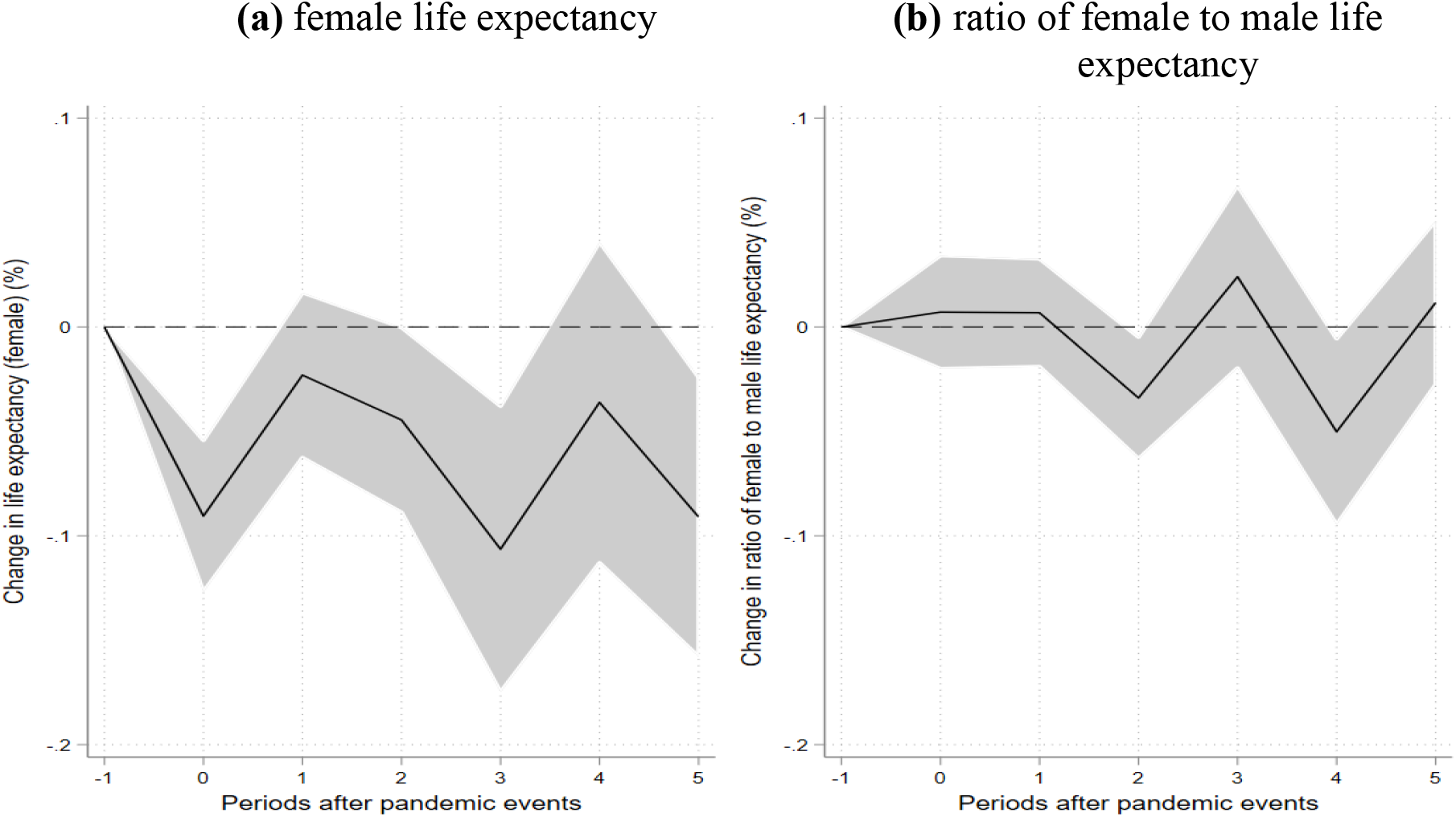
The response of life expectancy indices to the shock from pandemic episodes. *Note*: Estimates of IFRs based on LPs (see equation 1). The dependent variable is the change in the logarithm of the female life expectancy (left panel) or change in the ratio of female to male life expectancy (right panel). See section 3.1 for a description of the model specification. Shaded areas are 90% confidence intervals. The horizontal axis shows years (*k*) after the start of the pandemic event with *k* = 0 denoting the year when the pandemic started. The sample covers 186 countries over 1960-2018 period.

We also considered other measures of gender and health inequality including indicators based on labour market participation rates, mortality, child mortality and total fertility rates (results not reported). In neither case were the effects found to be statistically significant.

## 5. Conclusions

This paper provides evidence on the short and medium-term impact of the six past major pandemics on income, wealth and gender inequalities. We confirmed findings of other studies that past pandemics had statistically significant, but a rather moderate impact on income inequality. On the other hand, we did not find evidence that wealth inequalities have been worsening by the pandemics. Our results concerning gender inequality are not fully consistent across measures of gender inequality, but we found some evidence of declining gender equality among the hardest hit countries, as well as of growing gender gaps in unemployment over the four years after the onset of the pandemic. Admittedly, the present paper is only a preliminary study on the impact of past pandemics on wealth and gender inequalities. The data for those two types of inequalities are still available only for a limited number of, mostly rich, countries and periods. Therefore, we cannot rule out that some of our results are affected by sample selection. On the other hand, the paper seems to provide a useful starting point for future analyses based on more comprehensive data.

## Data Availability

The data are available upon request.

## Supplementary Appendix – for online publication only

**Table A1.** List of pandemic events.

| Announcement year | Event name | Affected countries (ISO codes) | Number of affected countries |
| --- | --- | --- | --- |
| 1968 | H3N2 Flu | ARG, AUS, CHL, DNK, FIN, FRA, GBR, GRC, HKG, ITA, JAM, JPN, NLD, NOR, PRT, SWE, USA, ZAF | 18 |
| 2003 | SARS | AUS, CAN, CHE, CHN, DEU, ESP, FRA, GBR, HKG, IDN, IND, IRL, ITA, KOR, KWT, MAC, MNG, MYS, NZL, PHL, ROU, RUS, SGP, SWE, THA, TWN, USA, VNM, ZAF | 29 |
| 2009 | H1N1 | AFG, AGO, ALB, AND, ARE, ARG, ARM, ATG, AUS, AUT, AZE, BDI, BEL, BGD, BGR, BHR, BHS, BIH, BLR, BLZ, BMU, BOL, BRA, BRB, BRN, BTN, BWA, CAN, CHE, CHL, CHN, CIV, CMR, COD, COG, COL, CPV, CRI, CUB, CYM, CYP, CZE, DEU, DJI, DMA, DNK, DOM, DZA, ECU, EGY, ESP, EST, ETH, FIN, FJI, FRA, FSM, GAB, GBR, GEO, GHA, GRC, GRD, GTM, GUY, HND, HRV, HTI, HUN, IDN, IND, IRL, IRN, IRQ, ISL, ISR, ITA, JAM, JOR, JPN, KAZ, KEN, KHM, KIR, KNA, KOR, KWT, LAO, LBN, LBY, LCA, LIE, LKA, LSO, LTU, LUX, LVA, MAR, MDA, MDG, MDV, MEX, MHL, MKD, MLI, MLT, MMR, MNE, MNG, MOZ, MUS, MWI, MYS, NAM, NGA, NIC, NLD, NOR, NPL, NRU, NZL, OMN, PAK, PAN, PER, PHL, PLW, PNG, POL, PRI, PRT, PRY, QAT, ROU, RUS, RWA, SAU, SDN, SGP, SLB, SLV, SOM, SRB, STP, SUR, SVK, SVN, SWE, SWZ, SYC, SYR, TCD, THA, TJK, TON, TTO, TUN, TUR, TUV, TZA, UGA, UKR, URY, USA, VCT, VEN, VNM, VUT, WSM, YEM, ZAF, ZMB, ZWE | 173 |
| 2012 | MERS | ARE, AUT, CHN, DEU, DZA, EGY, FRA, GBR, GRC, IRN, ITA, JOR, KOR, KWT, LBN, MYS, NLD, OMN, PHL, QAT, SAU, THA, TUN, TUR, USA, YEM | 26 |
| 2014 | Ebola | ESP, GBR, GIN, ITA, LBR, MLI, NGA, SEN, SLE, USA | 10 |
| 2016 | Zika | ABW, ARG, ATG, BHS, BLZ, BOL, BRA, BRB, CAN, CHL, COL, CRI, CUB, CYM, DMA, DOM, ECU, GRD, GTM, GUY, HND, HTI, JAM, KNA, LCA, NIC, PAN, PER, PRI, PRY, SLV, SUR, TCA, TTO, URY, USA, VCT, VEN | 38 |

Source: Ma et al. (2020) and own construction.

**Table A2.** Summary statistics.

|  | Mean | Standard deviation | Minimum | Maximum | N |
| --- | --- | --- | --- | --- | --- |
| Top 10% income share | 42.28 | 10.45 | 15.33 | 79.87 | 5651 |
| Top 1% income share | 14.48 | 5.99 | 1.94 | 63.76 | 5752 |
| Gini index | 52.46 | 10.28 | 12.37 | 83.27 | 5383 |
| Top 10% wealth share | 62.89 | 8.59 | 34.70 | 89.00 | 884 |
| Top 1% wealth share | 32.16 | 9.80 | 10.60 | 74.50 | 884 |
| Gender Inequality Index (GII) | 0.38 | 0.19 | 0.03 | 0.82 | 1943 |
| Global Gender Gap Index (GGGI) | 0.69 | 0.06 | 0.52 | 0.88 | 1839 |
| Female labor force participation rate | 55.82 | 17.47 | 5.94 | 91.83 | 5797 |
| Ratio of female to male labor force participation rate (x 100) | 71.18 | 20.97 | 8.26 | 109.12 | 5797 |
| Female unemployment rate | 9.25 | 7.51 | 0.12 | 47.65 | 5610 |
| Ratio of female to male unemployment rate (x 100) | 137.87 | 129.46 | 11.23 | 2886.67 | 5610 |
| Female life expectancy (female) | 66.54 | 12.05 | 22.39 | 87.70 | 11531 |
| Ratio of female to male life expectancy (x 100) | 107.55 | 3.55 | 95.89 | 137.50 | 11531 |
| Female child (under-5) mortality rate, under-5, female | 71.03 | 73.89 | 1.50 | 410.70 | 10434 |
| Ratio of female to male child mortality (x 100) | 85.78 | 6.21 | 70.74 | 124.00 | 10434 |
| Female mortality rate | 191.69 | 119.52 | 24.49 | 796.06 | 11294 |
| Ratio of female to male mortality rate | 68.96 | 16.75 | 28.97 | 131.30 | 11294 |
| Total fertility rate | 4.00 | 2.02 | 0.86 | 8.86 | 11548 |
| Pandemic episodes dummy | 0.02 | 0.14 | 0.00 | 1.00 | 13685 |
| Real GDP per capita | 11901.74 | 18759.82 | 132.07 | 196061.42 | 9427 |
| Population density | 328.55 | 1583.75 | 0.10 | 21389.10 | 12164 |
| Urban population (% of total population) | 51.14 | 25.76 | 2.08 | 100.00 | 12832 |
| KOF Trade Globalisation Index | 49.08 | 17.74 | 9.01 | 96.03 | 8335 |
| KOF Financial Globalisation Index | 50.05 | 18.43 | 6.91 | 98.03 | 8407 |

Source: own construction. See section 3 in the main text for the description of data sources.

**Table A3.** Impact of pandemics on pre-tax top 1% income share.

|  | k=0 | k=1 | k=2 | k=3 | k=4 | k=5 |
| --- | --- | --- | --- | --- | --- | --- |
| $D_{i,t}$ | 0.005<br>(0.008) | -0.007<br>(0.008) | 0.026***<br>(0.008) | 0.015**<br>(0.007) | 0.032***<br>(0.010) | 0.018*<br>(0.010) |
| $D_{i,t-1}$ | -0.013*<br>(0.007) | 0.009<br>(0.009) | 0.000<br>(0.008) | 0.016*<br>(0.010) | 0.005<br>(0.009) | -0.007<br>(0.010) |
| $D_{i,t-2}$ | 0.020***<br>(0.006) | 0.016**<br>(0.006) | 0.024***<br>(0.009) | 0.014<br>(0.009) | 0.001<br>(0.009) | 0.012<br>(0.011) |
| $\Delta I_{i,t-1}$ | -0.006<br>(0.061) | 0.695***<br>(0.048) | 0.790***<br>(0.078) | 0.869***<br>(0.079) | 0.930***<br>(0.064) | 0.900***<br>(0.092) |
| $\Delta I_{i,t-2}$ | 0.064**<br>(0.032) | -0.315***<br>(0.028) | -0.187**<br>(0.072) | -0.176**<br>(0.072) | -0.181***<br>(0.060) | -0.115<br>(0.085) |
| Constant | -0.023**<br>(0.011) | -0.015<br>(0.023) | -0.011<br>(0.028) | -0.014<br>(0.012) | -0.029<br>(0.028) | -0.026<br>(0.026) |
| N | 5030 | 4853 | 4677 | 4510 | 4343 | 4176 |
| R<sup>2</sup> | 0.089 | 0.441 | 0.563 | 0.662 | 0.735 | 0.768 |

Note: results based on the OLS estimation of equation (1) using a sample of 167 countries observed over 1950-2019. See section 3.2 for a description of data sources. Standard errors in parentheses clustered at the country level. \* $p < 0.1$ , \*\* $p < 0.05$ , \*\*\* $p < 0.01$ . Country and time fixed effects are included but not reported.

**Table A4.** Impact of pandemics on pre-tax Gini coefficient for income inequality.

|  | k=0 | k=1 | k=2 | k=3 | k=4 | k=5 |
| --- | --- | --- | --- | --- | --- | --- |
| $D_{i,t}$ | 0.002<br>(0.002) | -0.000<br>(0.002) | 0.007***<br>(0.003) | 0.003<br>(0.003) | 0.012***<br>(0.004) | 0.003<br>(0.004) |
| $D_{i,t-1}$ | -0.002<br>(0.002) | 0.002<br>(0.002) | -0.001<br>(0.003) | 0.005*<br>(0.003) | -0.003<br>(0.004) | -0.001<br>(0.005) |
| $D_{i,t-2}$ | 0.004**<br>(0.002) | 0.002<br>(0.003) | 0.010***<br>(0.003) | 0.002<br>(0.004) | 0.006<br>(0.004) | 0.004<br>(0.004) |
| $\Delta I_{i,t-1}$ | 0.069<br>(0.060) | 0.774***<br>(0.051) | 0.825***<br>(0.077) | 0.942***<br>(0.075) | 0.981***<br>(0.085) | 1.001***<br>(0.090) |
| $\Delta I_{i,t-2}$ | 0.153***<br>(0.036) | -0.285***<br>(0.026) | -0.171**<br>(0.066) | -0.212***<br>(0.067) | -0.210**<br>(0.082) | -0.197**<br>(0.087) |
| Constant | 0.006<br>(0.007) | -0.013<br>(0.014) | -0.018*<br>(0.009) | -0.013**<br>(0.006) | -0.027<br>(0.019) | 0.014<br>(0.036) |
| $N$ | 4745 | 4575 | 4406 | 4238 | 4070 | 3901 |
| $R^2$ | 0.132 | 0.522 | 0.642 | 0.737 | 0.793 | 0.831 |

Note: see note to Table A3.

**Table A5.** Impact of pandemics on top 10% wealth share.

|  | k=0 | k=1 | k=2 | k=3 | k=4 | k=5 |
| --- | --- | --- | --- | --- | --- | --- |
| $D_{i,t}$ | -0.002<br>(0.003) | -0.002<br>(0.004) | -0.000<br>(0.006) | 0.004<br>(0.006) | 0.008<br>(0.008) | 0.003<br>(0.008) |
| $D_{i,t-1}$ | -0.001<br>(0.004) | 0.001<br>(0.007) | 0.006<br>(0.005) | 0.009<br>(0.008) | 0.004<br>(0.008) | -0.012<br>(0.010) |
| $D_{i,t-2}$ | 0.000<br>(0.006) | 0.005<br>(0.005) | 0.010<br>(0.008) | 0.006<br>(0.008) | -0.011<br>(0.009) | 0.006<br>(0.012) |
| $\Delta I_{i,t-1}$ | -0.247***<br>(0.074) | 0.539***<br>(0.067) | 0.716***<br>(0.091) | 0.694***<br>(0.070) | 0.716***<br>(0.073) | 0.696***<br>(0.067) |
| $\Delta I_{i,t-2}$ | -0.220**<br>(0.082) | -0.487***<br>(0.085) | -0.217*<br>(0.115) | -0.002<br>(0.083) | 0.016<br>(0.086) | 0.022<br>(0.080) |
| Constant | -0.005*<br>(0.002) | -0.007**<br>(0.003) | -0.006*<br>(0.004) | -0.011***<br>(0.004) | -0.015***<br>(0.004) | -0.018***<br>(0.005) |
| $N$ | 736 | 689 | 643 | 597 | 551 | 505 |
| $R^2$ | 0.200 | 0.357 | 0.517 | 0.619 | 0.691 | 0.715 |

Note: see note to Table A3.

**Table A6.** Impact of pandemics on top 1% wealth share.

|  | k=0 | k=1 | k=2 | k=3 | k=4 | k=5 |
| --- | --- | --- | --- | --- | --- | --- |
| $D_{i,t}$ | 0.001<br>(0.007) | -0.010<br>(0.009) | 0.005<br>(0.015) | 0.006<br>(0.011) | -0.003<br>(0.015) | 0.002<br>(0.019) |
| $D_{i,t-1}$ | -0.010<br>(0.009) | 0.002<br>(0.016) | 0.004<br>(0.011) | -0.001<br>(0.015) | 0.001<br>(0.018) | -0.027<br>(0.020) |
| $D_{i,t-2}$ | 0.003<br>(0.015) | 0.013<br>(0.010) | 0.004<br>(0.014) | 0.008<br>(0.019) | -0.026<br>(0.020) | 0.008<br>(0.020) |
| $\Delta I_{i,t-1}$ | -0.305***<br>(0.084) | 0.539***<br>(0.077) | 0.651***<br>(0.103) | 0.623***<br>(0.092) | 0.668***<br>(0.083) | 0.654***<br>(0.085) |
| $\Delta I_{i,t-2}$ | -0.174**<br>(0.083) | -0.460***<br>(0.097) | -0.106<br>(0.128) | 0.077<br>(0.099) | 0.058<br>(0.088) | 0.053<br>(0.095) |
| Constant | -0.017***<br>(0.006) | -0.016**<br>(0.007) | -0.024**<br>(0.009) | -0.029***<br>(0.007) | -0.032***<br>(0.009) | -0.045***<br>(0.012) |
| $N$ | 736 | 689 | 643 | 597 | 551 | 505 |
| $R^2$ | 0.216 | 0.379 | 0.547 | 0.632 | 0.708 | 0.732 |

Note: see note to Table A3.

**Table A7.**
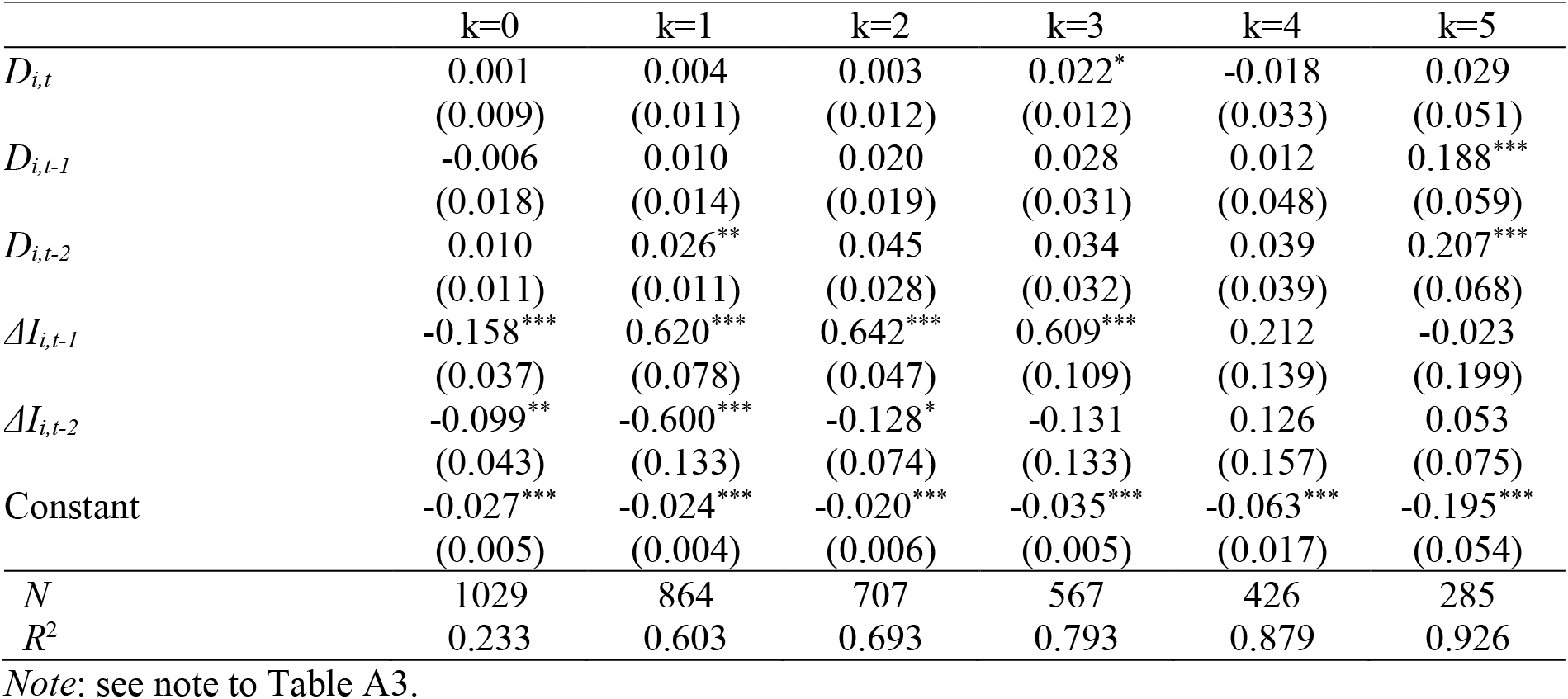
Impact of pandemics on Gender Inequality Index (GII)

|  | k=0 | k=1 | k=2 | k=3 | k=4 | k=5 |
| --- | --- | --- | --- | --- | --- | --- |
| $D_{i,t}$ | 0.001<br>(0.009) | 0.004<br>(0.011) | 0.003<br>(0.012) | 0.022*<br>(0.012) | -0.018<br>(0.033) | 0.029<br>(0.051) |
| $D_{i,t-1}$ | -0.006<br>(0.018) | 0.010<br>(0.014) | 0.020<br>(0.019) | 0.028<br>(0.031) | 0.012<br>(0.048) | 0.188***<br>(0.059) |
| $D_{i,t-2}$ | 0.010<br>(0.011) | 0.026**<br>(0.011) | 0.045<br>(0.028) | 0.034<br>(0.032) | 0.039<br>(0.039) | 0.207***<br>(0.068) |
| $\Delta I_{i,t-1}$ | -0.158***<br>(0.037) | 0.620***<br>(0.078) | 0.642***<br>(0.047) | 0.609***<br>(0.109) | 0.212<br>(0.139) | -0.023<br>(0.199) |
| $\Delta I_{i,t-2}$ | -0.099**<br>(0.043) | -0.600***<br>(0.133) | -0.128*<br>(0.074) | -0.131<br>(0.133) | 0.126<br>(0.157) | 0.053<br>(0.075) |
| Constant | -0.027***<br>(0.005) | -0.024***<br>(0.004) | -0.020***<br>(0.006) | -0.035***<br>(0.005) | -0.063***<br>(0.017) | -0.195***<br>(0.054) |
| $N$ | 1029 | 864 | 707 | 567 | 426 | 285 |
| $R^2$ | 0.233 | 0.603 | 0.693 | 0.793 | 0.879 | 0.926 |

Note: see note to Table A3.

**Table A8.** Impact of pandemics on Global Gender Gap Index (GGGI)

|  | k=0 | k=1 | k=2 | k=3 | k=4 | k=5 |
| --- | --- | --- | --- | --- | --- | --- |
| $D_{i,t}$ | -0.001<br>(0.002) | 0.008**<br>(0.004) | 0.001<br>(0.004) | -0.005<br>(0.005) | -0.007<br>(0.006) | -0.010*<br>(0.006) |
| $D_{i,t-1}$ | 0.007**<br>(0.003) | 0.002<br>(0.003) | 0.001<br>(0.005) | 0.002<br>(0.006) | -0.006<br>(0.007) | -0.000<br>(0.006) |
| $D_{i,t-2}$ | -0.001<br>(0.003) | -0.005<br>(0.004) | -0.004<br>(0.005) | -0.007<br>(0.006) | -0.002<br>(0.004) | -0.011*<br>(0.006) |
| $\Delta I_{i,t-1}$ | -0.152***<br>(0.034) | 0.470***<br>(0.033) | 0.613***<br>(0.045) | 0.584***<br>(0.057) | 0.594***<br>(0.048) | 0.420***<br>(0.067) |
| $\Delta I_{i,t-2}$ | -0.148***<br>(0.034) | -0.420***<br>(0.029) | -0.270***<br>(0.047) | -0.154***<br>(0.048) | -0.101**<br>(0.043) | -0.060<br>(0.054) |
| Constant | 0.025***<br>(0.002) | 0.011***<br>(0.004) | 0.009**<br>(0.004) | 0.002<br>(0.006) | -0.018***<br>(0.007) | -0.012*<br>(0.006) |
| $N$ | 1235 | 1091 | 950 | 819 | 697 | 577 |
| $R^2$ | 0.143 | 0.406 | 0.487 | 0.565 | 0.677 | 0.728 |

Note: see note to Table A3.

**Table A9.** Impact of pandemics on female unemployment rate.

|  | k=0 | k=1 | k=2 | k=3 | k=4 | k=5 |
| --- | --- | --- | --- | --- | --- | --- |
| $D_{i,t}$ | 0.037***<br>(0.014) | 0.012<br>(0.013) | 0.028<br>(0.020) | 0.009<br>(0.016) | 0.014<br>(0.015) | -0.007<br>(0.020) |
| $D_{i,t-1}$ | 0.005<br>(0.012) | 0.016<br>(0.018) | -0.005<br>(0.013) | -0.001<br>(0.013) | -0.014<br>(0.018) | -0.002<br>(0.021) |
| $D_{i,t-2}$ | 0.023<br>(0.016) | 0.024*<br>(0.012) | 0.027**<br>(0.013) | 0.001<br>(0.018) | 0.016<br>(0.019) | -0.055***<br>(0.021) |
| $\Delta I_{i,t-1}$ | 0.114***<br>(0.035) | 0.763***<br>(0.033) | 0.868***<br>(0.047) | 0.964***<br>(0.046) | 1.010***<br>(0.044) | 1.020***<br>(0.048) |
| $\Delta I_{i,t-2}$ | 0.060**<br>(0.026) | -0.321***<br>(0.030) | -0.260***<br>(0.045) | -0.281***<br>(0.042) | -0.275***<br>(0.037) | -0.254***<br>(0.042) |
| Constant | 0.045***<br>(0.016) | 0.023<br>(0.015) | 0.014<br>(0.017) | -0.022<br>(0.016) | 0.007<br>(0.019) | -0.020<br>(0.018) |
| $N$ | 4805 | 4618 | 4441 | 4264 | 4087 | 3909 |
| $R^2$ | 0.135 | 0.522 | 0.625 | 0.708 | 0.754 | 0.789 |

Note: see note to Table A3.

**Table A10.** Impact of pandemics on the ratio of female to male unemployment rate.

|  | k=0 | k=1 | k=2 | k=3 | k=4 | k=5 |
| --- | --- | --- | --- | --- | --- | --- |
| $D_{i,t}$ | -0.008<br>(0.012) | 0.013<br>(0.011) | 0.019<br>(0.012) | 0.018<br>(0.014) | 0.037***<br>(0.014) | -0.006<br>(0.017) |
| $D_{i,t-1}$ | 0.000<br>(0.009) | -0.006<br>(0.011) | 0.000<br>(0.011) | 0.016<br>(0.015) | -0.015<br>(0.018) | 0.000<br>(0.019) |
| $D_{i,t-2}$ | 0.002<br>(0.009) | -0.001<br>(0.011) | 0.017<br>(0.013) | -0.022<br>(0.018) | -0.006<br>(0.017) | -0.015<br>(0.014) |
| $\Delta I_{i,t-1}$ | -0.129***<br>(0.038) | 0.583***<br>(0.032) | 0.651***<br>(0.048) | 0.712***<br>(0.046) | 0.782***<br>(0.045) | 0.729***<br>(0.057) |
| $\Delta I_{i,t-2}$ | 0.005<br>(0.023) | -0.323***<br>(0.024) | -0.130***<br>(0.042) | -0.091**<br>(0.039) | -0.093**<br>(0.036) | 0.001<br>(0.048) |
| Constant | 0.016*<br>(0.009) | 0.021**<br>(0.009) | 0.022**<br>(0.010) | 0.023**<br>(0.009) | 0.021*<br>(0.011) | -0.001<br>(0.010) |
| $N$ | 4805 | 4618 | 4441 | 4264 | 4087 | 3909 |
| $R^2$ | 0.067 | 0.353 | 0.433 | 0.535 | 0.629 | 0.665 |

Note: see note to Table A3.

**Table A11.** Impact of pandemics on the female life expectancy.

|  | k=0 | k=1 | k=2 | k=3 | k=4 | k=5 |
| --- | --- | --- | --- | --- | --- | --- |
| $D_{i,t}$ | -0.001***<br>(0.000) | -0.000<br>(0.000) | -0.000<br>(0.000) | -0.001**<br>(0.001) | -0.000<br>(0.001) | -0.001*<br>(0.001) |
| $D_{i,t-1}$ | 0.000<br>(0.000) | 0.001**<br>(0.000) | -0.000<br>(0.000) | 0.001<br>(0.001) | -0.000<br>(0.001) | 0.001<br>(0.001) |
| $D_{i,t-2}$ | 0.000<br>(0.000) | -0.001**<br>(0.000) | -0.000<br>(0.001) | -0.002**<br>(0.001) | -0.001<br>(0.000) | -0.001<br>(0.001) |
| $\Delta I_{i,t-1}$ | 0.698***<br>(0.203) | 1.422***<br>(0.160) | 1.578***<br>(0.133) | 1.665***<br>(0.091) | 1.718***<br>(0.077) | 1.762***<br>(0.056) |
| $\Delta I_{i,t-2}$ | 0.132<br>(0.189) | -0.554***<br>(0.164) | -0.687***<br>(0.144) | -0.763***<br>(0.103) | -0.807***<br>(0.088) | -0.841***<br>(0.066) |
| Constant | 0.002***<br>(0.000) | 0.003***<br>(0.000) | 0.002***<br>(0.000) | 0.004***<br>(0.001) | 0.003***<br>(0.001) | 0.003***<br>(0.001) |
| $N$ | 10274 | 10089 | 9904 | 9718 | 9532 | 9346 |
| $R^2$ | 0.716 | 0.909 | 0.947 | 0.964 | 0.974 | 0.981 |

Note: see note to Table A3.

**Table A12.** Impact of pandemics on the ratio of female to male life expectancy.

|  | k=0 | k=1 | k=2 | k=3 | k=4 | k=5 |
| --- | --- | --- | --- | --- | --- | --- |
| $D_{i,t}$ | 0.000<br>(0.000) | 0.000<br>(0.000) | -0.000<br>(0.000) | 0.000<br>(0.000) | -0.001<br>(0.000) | 0.000<br>(0.000) |
| $D_{i,t-1}$ | 0.000<br>(0.000) | -0.000<br>(0.000) | 0.000<br>(0.000) | -0.001*<br>(0.000) | -0.000<br>(0.000) | -0.000<br>(0.000) |
| $D_{i,t-2}$ | -0.000<br>(0.000) | 0.000<br>(0.000) | -0.000<br>(0.000) | -0.000<br>(0.000) | 0.000<br>(0.000) | 0.000<br>(0.000) |
| $\Delta I_{i,t-1}$ | 0.152<br>(0.157) | 0.848***<br>(0.159) | 0.892***<br>(0.263) | 1.070***<br>(0.228) | 1.151***<br>(0.250) | 1.269***<br>(0.219) |
| $\Delta I_{i,t-2}$ | 0.314***<br>(0.060) | -0.165*<br>(0.094) | -0.087<br>(0.240) | -0.225<br>(0.212) | -0.281<br>(0.241) | -0.381*<br>(0.213) |
| Constant | 0.000*<br>(0.000) | 0.000**<br>(0.000) | 0.000**<br>(0.000) | 0.000**<br>(0.000) | 0.001***<br>(0.000) | 0.001***<br>(0.000) |
| $N$ | 10274 | 10089 | 9904 | 9718 | 9532 | 9346 |
| $R^2$ | 0.183 | 0.575 | 0.703 | 0.800 | 0.838 | 0.882 |

Note: see note to Table A3.

## Footnotes

1 Aburto et al. (2020) analyse the impact of HIV/AIDS epidemic on another dimension of inequalities – black-white disparities in life expectancy. Clay et al. (2019) study the disparities in excess mortality across 438 US cities during the 1918 Influenza Pandemic. Aparicio Fenoll and Grossbard (2020) show that cross-country fatalities from COVID-19 are associated with differences in intergenerational residence patterns. Markowitz et al. (2019) find that increases in the US unemployment rate lead to the spread of flu which can contribute to flu pandemic.

2 Specifically, we consider the distribution of household net worth understood as the sum of households’ financial and nonfinancial assets minus households’ debts.

3 Several papers have recently applied the LP to study the effects of past pandemics on various outcomes – see Furceri et al. (2020), Jordà et al, (2020), Ma et al. (2020) and S aadi-Sedik and Xu (2020).

4 In some specifications, we replace the pandemic dummy with pandemic severity dummies (see section 3.2.1) to account for the fact that different health crises hit some countries relatively lightly (with no or little excess mortality), while had very significant impact on other countries.

5 See Ma et al. (2020) for a detailed description of these pandemic episodes.

6 The pre-tax pre-transfer national income is defined as the sum of all income flows accruing to the individual owners of labour and capital before the operation of the tax and transfer system, but after the operation of the pension and unemployment insurance systems. The DINA provides also inequality indices in terms of the post-tax post-transfer national income, but these are available only for about 40 advanced countries.

7 The *Global wealth reports* data are available online at https://www.credit-suisse.com/about-us/en/reports-research/global-wealth-report.html. The database contains also the Gini coefficients for wealth distribution, but only since 2010. Recently, Islam and McGillivray (2020) used top wealth shares from *Global wealth report* to study the link between wealth inequality and economic growth.

8 Similar results were obtained for the inequality measured using the top 10% income shares.

9 Full results are available upon request.

10 We do not report these results to save space, but they are available upon request.

11 Results for groups of countries differentiated by pandemic severity are also statistically insignificant.

12 In case of the GII, the estimates by pandemic severity show small increases in gender inequality within the horizon of three years that fade away afterwards.

## Notes

### Competing Interest Statement

The authors have declared no competing interest.

### Funding Statement

There has been no funding for this research.

### Author Declarations

University of Warsaw body.

